# Women undergoing repeated bariatric surgery due to recurrent weight gain exhibit an inflammatory molecular and functional signature of subcutaneous adipose tissue

**DOI:** 10.64898/2026.05.30.26354509

**Authors:** Ariel Shneyour, Yuval G. Noach, Uri Yoel, Marina Rosengarten-Levin, Oleg Zilber, Alon Zemer, Habib Muallem, Vered Chalifa-Caspi, Danit R. Shahar, Idit F. Liberty, Nur Elkarnawi, Oleg Dukhno, Idan Carmeli, Ran Orgad, Yulia Haim, Assaf Rudich

**Author notes:** Co-corresponding authors: Yulia Haim, Ph.D., Department of Clinical Biochemistry and Pharmacology, Faculty of Health Sciences, Ben-Gurion University of the Negev, Beer-Sheva, Israel, Assaf Rudich, M.D., Ph.D., Department of Clinical Biochemistry and Pharmacology, Faculty of Health Sciences, Ben-Gurion University of the Negev, Beer-Sheva, Israel.

## Abstract

**Background:** Repeated metabolic-bariatric surgery (MBS, r-BS) represents 10–25% of all MBS procedures and is commonly performed for recurrent weight gain after initial weight loss. How weight loss followed by regain reshapes adipose tissue biology remains unclear. We hypothesized that women undergoing r-BS exhibit a distinct adipose tissue signature compared with those undergoing primary bariatric surgery (p-BS).

**Methods:** We analyzed subcutaneous and visceral adipose tissues (SAT, VAT, respectively) from women undergoing either p-BS, or r-BS with documented, >15% weight-loss after prior MBS. Tissues were assessed histologically, molecularly, and functionally (activation of human microglia cells (HMC3) by SAT secretome).

**Results:** Consistent with other cohorts, women undergoing r-BS (n=21) trended to be older (47.2 vs. 40.5 y, p=0.06) than those undergoing p-BS (n=35), with a lower BMI (42.3 vs. 45.6 kg/m2, respectively, p=0.103), and a trend for improved cardiometabolic risk parameters such as fasting insulin, CRP and HDL-c. Adipose tissues’ histological features (adipocyte size, fibrosis, macrophage and crown-like structures abundance) were similar, while adipose mast-cells were slightly (though insignificantly) more prevalent in r-BS. Single-nucleus RNA-seq-based deconvolution algorithm applied unto bulk RNA-seq to uncover differences in cell composition confirmed the absence of a major shift in adipose tissue cell-type composition. Yet, it uncovered unique transcriptome of SAT, with activation of inflammatory pathways in r-BS. Consistently, SAT explants from r-BS secreted higher protein concentrations of NFκB-regulated cytokines IL6 and IL8. Biological impact of the more inflammatory secretome was demonstrated by its increased ability to inflammatory activate human microglia cells.

**Conclusions:** Prior BS with significant weight-loss-regain in women is associated with inflammatory transcriptome and secretome of SAT, possibly reflecting on adipose-brain endocrine communication.

## Introduction

Despite unprecedented advances in obesity management medications, bariatric surgery remains the most proven to be both effective and durable treatment for severe obesity and its complications ^1–3^. As the prevalence of obesity continues to rise worldwide, the number of metabolic bariatric procedures (MBS) performed annually has increased substantially ^4^. This suggests MBS will remain an integral component of contemporary management particularly for severe obesity.

Despite its effectiveness, some patients experience suboptimal clinical response and/or recurrent weight gain following an initial significant weight loss ^5,6^. In such cases, a repeated MBS may be considered to restore weight loss by revising (intensifying) the previous procedure or conversion to a different MBS with new mechanism of action ^7^. The number of repeated procedures has increased steadily ^8^, and currently may reach ~25% of MBS. Compared with primary operations, repeated procedures are technically more complex and are generally associated with longer operative times and higher complication rates ^9,10^. In addition, the clinical responses to repeated surgery are often smaller compared to primary MBS and display greater variability in weight loss outcomes ^11^. Before surgery, r-BS candidates were reported to clinically present with lower body mass index (BMI) compared to p-BS candidates, and a longer history of severe obesity and interventions. Thus, they are likely to be older, but to exhibit pre-surgical trends towards better cardio-metabolic risk parameters. These clinical characteristics could reflect selection bias, and/or be partially attributable to their somewhat lower BMI compared to p-BS candidates. But in addition, this complex clinical scenario also raises the possibility of prior-MBS-induced significant weight loss and its lasting beneficial effects despite subsequent recurrent weight gain, as recently shown in response to life-style interventions ^12^. Although “obesogenic memory” and some molecular mechanisms that may encode it have been described even in humans ^13^, the context is usually in the extended impact of obesity into the post-obese lean state. Understanding the possible impact of prior MBS-induced significant weight loss and recurrent weight gain (weight cycling), at the tissue and molecular level is still lacking.

Adipose tissue(s)’ adaptation to obesity is strongly associated with, and was implicated in, the tendency to develop obesity-associated medical problems ^14^. Adipose tissues undergo extensive remodeling characterized by immune cell infiltration ^15^, fibrosis ^16^, and altered secretory profile ^17,18^, which are thought to contribute to systemic inflammation and metabolic dysfunction ^19^. While clinically r-BS differ from p-BS candidates, tissue and molecular-level comparisons, that can deepen mechanistic understanding of their unique characteristics and clinical course, are scarce. Here we hypothesized that candidates of r-BS present distinct adipose tissue signatures compared to p-BS candidates. To challenge our hypothesis, we performed a multi-level characterization of adipose tissue obtained during MBS, that included systematic histological assessment, transcriptomic profiling, and functional analyses of adipose tissue-derived secretory activity.

## Methods

### Study participants and clinical data collection

Patients undergoing MBS at Soroka University Medical Center (Be’er-Sheva, Israel) provided in advance written informed consent for the collection and storage of blood and adipose tissue (AT) samples (visceral/omental and abdominal subcutaneous) to a biobank through which histological and molecular analyses studies were previously performed ^20–22^. Inclusion criteria were age 18–65 years, and obesity was defined as body mass index (BMI)≥30 kg/m^2^. Patients with known malignancies or acute illness/trauma were excluded. The study protocol was approved by the Helsinki Ethics Committee of Soroka University Medical Center (approval number: 15-0348). In the current study, we included female patients (women living with obesity) referred for MBS between 2022 and 2024. Participants were included in the primary MBS (p-BS) group if undergoing MBS for the first time. Repeated MBS (r-BS) included women who had: i. a history of a prior bariatric procedure; ii. a documented ≥15% weight loss following that procedure, which we reasoned was more likely to differentiated MBS-induced weight loss from other (often undocumented) intentional weight losses; iii. Documented subsequent weight regain. Clinical parameters, including BMI, lipid profile (total cholesterol, low-density lipoprotein [LDL], high-density lipoprotein cholesterol [HDL-c], and triglycerides [TG]), and glycemic measures (fasting glucose, insulin, and hemoglobin A1c [HbA1c] and the corresponding calculated Homeostatic Model Assessment for Insulin Resistance [HOMA-IR]) were obtained from the medical records when available or measured preoperatively.

### Histology

Biopsies of visceral adipose tissue (VAT) were sampled from the greater omentum and subcutaneous samples (SAT) from abdominal subcutaneous fat tissue, fixed in fresh 4% formalin for at least 24 h and embedded in paraffin. All following procedures were performed in the Clinical Pathology Laboratories of Maccabi Health Services (Rehovot, Israel), operating under ISO 1589:2022 standards, using standard clinical pathology laboratory procedures. In brief, paraffin blocks were sectioned (4□μm) and stained as follows: i. adipocyte size estimation: hematoxylin and eosin (H&E) for adipocyte size estimation (of at least 100 adipocytes per sample); ii. Total fibrosis estimation: Picrosirius Red solution (0.1% Sirius Red F3B in picric acid; SigmalZIAldrich, St. Louis, MO, USA). Iii. Macrophages: immunohistochemistry (IHC) staining was performed with the BenchMark ULTRA system (Roche, Tucson, AZ, USA) using anti-CD68 (Kp-1 Mouse monoclonal antibody, Cell Marque, Netherlands). CD68-positive cells were identified in at least 8 regions of interest of 500 µm^2^ and their abundance presented per 100 adipocytes. iv. Crown-like structures (CLS) were identified manually as adipocyte/lipid droplet surrounded by CD68-positive cells (macrophages) along >50% of its perimeter. V. Mast cells were identified by c-kit IHC staining using Polyclonal Rabbit Anti-Human CD117 antibody (DAKO, Agilent Technologies, Santa Clara, CA, USA). Stained slides were scanned on NanoZoomer S360 Digital slide scanner (Hamamatsu Photonics, Hamamatsu, Japan), and images were analyzed with an open-source image analysis software QuPath version 0.4.4.

### RNA extraction and bulk RNA-sequencing analysis

Total RNA from 19 samples of both visceral and subcutaneous adipose tissue biopsies (10 p-BS and 9 r-BS) was extracted using RNeasy Lipid Tissue Mini Kit (QIAGEN, Germantown, MD). RNA integrity was determined on a QIAxcel device (QIAGEN, QIAxcel RNA QC Kit v2.0), included were 19 samples with best RIN score (all>7), and concentration was determined using a QuantiFluor® RNA System (Promega, #E3310) on a Qubit™ Flex Fluorometer (Invitrogen). RNA-seq libraries were generated from polyA-enriched (NEBNext® Poly(A) mRNA Magnetic Isolation Module, NEB #E7490L) purified RNA using the recommended protocol (NEBNext® Ultra™ II RNA Library Prep Kit for Illumina®, NEB #E7775). The molarity of libraries was determined using QIAxcel (QIAxcel DNA High Sensitivity Kit) and QuantiFluor® dsDNA System (Promega, # E2670) on a Qubit™ Flex Fluorometer (Invitrogen). Libraries were sequenced (150PE) on a NovaSeq X (Illumina) in two independent, balanced experimental batches. Raw sequencing reads were processed using the NeatSeq-Flow platform (https://neatseq-flow.readthedocs.io/en/latest/). Quality control was performed using FastQC and MultiQC, and reads were trimmed using Trim Galore and cutadapt. Filtered reads were aligned to the human reference genome (GRCh38) using Bowtie2, and gene-level counts were generated using RSEM. Differential gene expression analysis was performed in R using the DESeq2 package ^23^. For comparison between study groups (p-BS vs. r-BS), the statistical model considered two effects: the study group and the technical batch. For each gene, log2 fold change, raw P value, and false discovery rate (FDR)-adjusted P value were calculated. Genes with an adjusted P value<0.05 were considered differentially expressed. For visualization of global transcriptomic patterns, surrogate variable analysis (SVA) was applied for batch correction prior to principal component analysis (PCA). Functional enrichment analysis was performed using the clusterProfiler package, focusing on KEGG pathways. Both over-representation analysis (ORA) and gene set enrichment analysis (GSEA) were applied; ORA was used to test enrichment among differentially expressed genes, while GSEA was used to assess enrichment across the full ranked gene list without applying a significance threshold. ChEA transcription factor enrichment analysis was performed on upregulated DEGs using the Enrichr platform ^24^.

### Deconvolution analysis

Cell-type composition was estimated using sNucConv ^25^, a deep-learning based deconvolution model trained on single-nucleus RNA sequencing data from human subcutaneous adipose tissue. The model adjusts for correlations between bulk RNA sequencing and single-nucleus data and outputs estimated proportions of adipocytes, adipose progenitor cells, immune cells, and vascular-associated cell types.

### Generation and classification of SAT condition medium

Human adipose tissue biopsies were freshly obtained (processed within 1 h of excision) from the operating room and washed twice with pre-warmed medium (MEM-α supplemented with 10% fetal bovine serum and 1% antibiotic–antimycotic solution. L-glutamine (2%) was added immediately prior to use). Samples were weighed, transferred to 12-well plates, and minced into ~2–3 mm^3^ fragments using sterile scissors and cultured (100 mg tissue per 1 mL medium) at 37 °C in a humidified atmosphere with 5% CO_2_. After 24 h incubation, medium was aspirated, and explants were washed once with fresh medium, and an additional 1 mL of fresh pre-warmed medium was added. “Conditioned medium” (AT-CM) was collected after this last 24 h incubation and stored at −80 °C until use. AT-CMs were categorized as being “low/high inflammatory” based on measurement of 3 cytokines (IL-6, IL-8 and CCL2) using specific Enzyme-Linked Immunosorbent Assay (ELISA, all from R&D Systems). Lowest/highest concentrations of secreted IL-6, IL-8 and CCL2 were categorized as low/high inflammatory range, respectively.

### Human microglia cell studies

Human microglia clone 3 (HMC3, American Type Culture Collection (ATCC), cell passages 3-6) were cultured in Eagle’s Minimum Essential Medium (EMEM) containing L-glutamine, supplemented with 10% fetal bovine serum and 2% antibiotic–antimycotic solution. Cells were seeded in 6-well plates and treated with 1 ml of LPS (1µg/ml) or AT-CM (1:5 dilution in EMEM), from either p-BS or r-BS groups, for 24 hours. Then, cells were washed and medium was replaced with fresh - 1% FBS EMEM culture media (800 µl per well) for additional 3.5h. Next, medium was collected and stored at – 80 °C until analysis. After medium collection, HMC3 were washed twice with ice cold PBS and frozen at – 80 °C. IL-6 secretion was measured using IL6 ELISA (R&D Systems) and normalized to total protein in cell lysates (Pierce™ BCA Protein Assay Kit (ThermoFisher Scientific)).

### Statistical analysis

Continuous variables are presented as mean (min-max), and categorical variables as n (%). Between-group comparisons (p-BS vs r-BS) were performed using the Mann-Whitney U test for continuous variables and Fisher’s exact test for categorical variables. Age-adjusted analyses were performed using regression models including group and age as predictors. Continuous variables were analyzed using linear regression, and categorical variables using logistic regression. All tests were two-tailed, and p values <0.05 were considered statistically significant. Analyses were performed in R (version 4.2.2) and GraphPad Prism (version 10.3.1).

## Results

Since a significant proportion of MBS are currently repeated procedures (revisional, conversion), we wished to better characterize this patient subgroup compared to those undergoing primary MBS (p-BS). For this, we recruited 56 women, 35 meeting criteria for p-BS and 21 for r-BS. All 21 women had undergone a previous adjustable gastric banding that induced a documented (in the medical records) ≥15% total body weight loss response following the procedure, with subsequent significant recurrent weight gain, and were recruited to undergo a conversion MBS. Clinical characteristics of the two groups were generally comparable, with no statistically significant differences across all parameters (**Table 1**). Yet, women in the r-BS tended to be older (47.2 vs. 40.5, p=0.06) and exhibited a more favourable metabolic profile consistent with previously reported cohorts ^9,10^, although none of these differences reached statistical significance, even after age-adjustment (**Table 1**).

**Table 1.** Cohort baseline clinical characteristics (N=56)

|  | **All**<br>(N=56 women) | **p-BS**<br>(N=35 women) | **r-BS**<br>(N=21 women) | **p value** | **Age-**<br>**adjusted**<br>**p value** |
| --- | --- | --- | --- | --- | --- |
| Age (years) | 43.0 (20.0, 68.0) | 40.5 (20.0, 68.0) | 47.2 (30.0, 62.0) | NS (0.060) |  |
| Above 50 years (%) | 17 (30.4%) | 10 (28.6%) | 7 (33.3%) | NS |  |
| BMI (kg/m<sup>2</sup>) | 44.4 (32.0, 59.2) | 45.6 (34.0, 59.2) | 42.4 (32.0, 57.0) | NS (0.077) | NS (0.116) |
| Type 2 DM (%) | 6 (10.7%) | 3 (8.6%) | 3 (14.3%) | NS | NS |
| SBP (mmHg) | 123.2 (88.0, 180.0) | 121.7 (88.0, 172.0) | 125.8 (101.0, 180.0) | NS | NS |
| DBP (mmHg) | 78.9 (58.0, 108.0) | 79.1 (60.0, 108.0) | 78.4 (58.0, 98.0) | NS | NS |
| Fasting glucose (mg/dL) | 91.3 (58.0, 133.0) | 91.9 (58.0, 153.0) | 90.2 (79.0, 114.0) | NS | NS |
| Fasting insulin (μU/ml) | 15.6 (2.9, 58.8) | 17.9 (2.9, 58.8) | 11.8 (4.6, 26.3) | NS (0.126) | NS (0.123) |
| HOMA-IR | 3.6 (0.8, 16.7) | 4.2 (0.8, 16.7) | 2.6 (0.9, 5.6) | NS | NS (0.124) |
| HbA1c (%) | 5.6 (4.4, 8.3) | 5.6 (4.6, 8.3) | 5.6 (4.4, 6.3) | NS | NS |
| Total Cholesterol (mg/dL) | 170.9 (108.0, 239.0) | 169.3 (108.0, 231.0) | 173.4 (129.0, 239.0) | NS | NS |
| Triglycerides (mg/dL) | 148.9 (52.0, 362.0) | 156.3 (52.0, 362.0) | 136.8 (61.0, 244.0) | NS | NS |
| LDL (mg/dL) | 97.6 (34.0, 157.0) | 95.7 (34.0, 157.0) | 100.7 (66.0, 153.0) | NS | NS |
| HDL-c (mg/dL) | 43.1 (31.0, 70.0) | 41.3 (31.0, 55.0) | 46.0 (34.0, 70.0) | NS (0.078) | NS (0.088) |
| hs-CRP (mg/dL) | 1.16 (0.14, 4.58) | 1.20 (0.18, 4.21) | 1.09 (0.14, 4.58) | NS (0.132) | NS |

p value < 0.15 is indicated in brackets.

We first assessed if adipose tissues (subcutaneous and visceral, SAT and VAT, respectively) significantly differed morphologically between the two groups, measuring histologically adipocyte size, total fibrosis, CD68-positive macrophage abundance, frequency of macrophage-based crown-like structures (CLS), and the number of mast (C-Kit (KIT)-positive) cells (**Figure 1, S1**). Both groups exhibited significant variability in each of these parameters, as shown in images of SAT samples from women in the minimal and maximal range from each group (**Figure 1**). Consequently, no statistically significant differences in the histo-pathological features of adipose tissue were detected, with only a non-significant trend towards higher relative abundance of mast cells in SAT, consistent with the better cardio-metabolic risk parameters of the r-BS group, as we previously reported ^26^.

**Figure 1.**
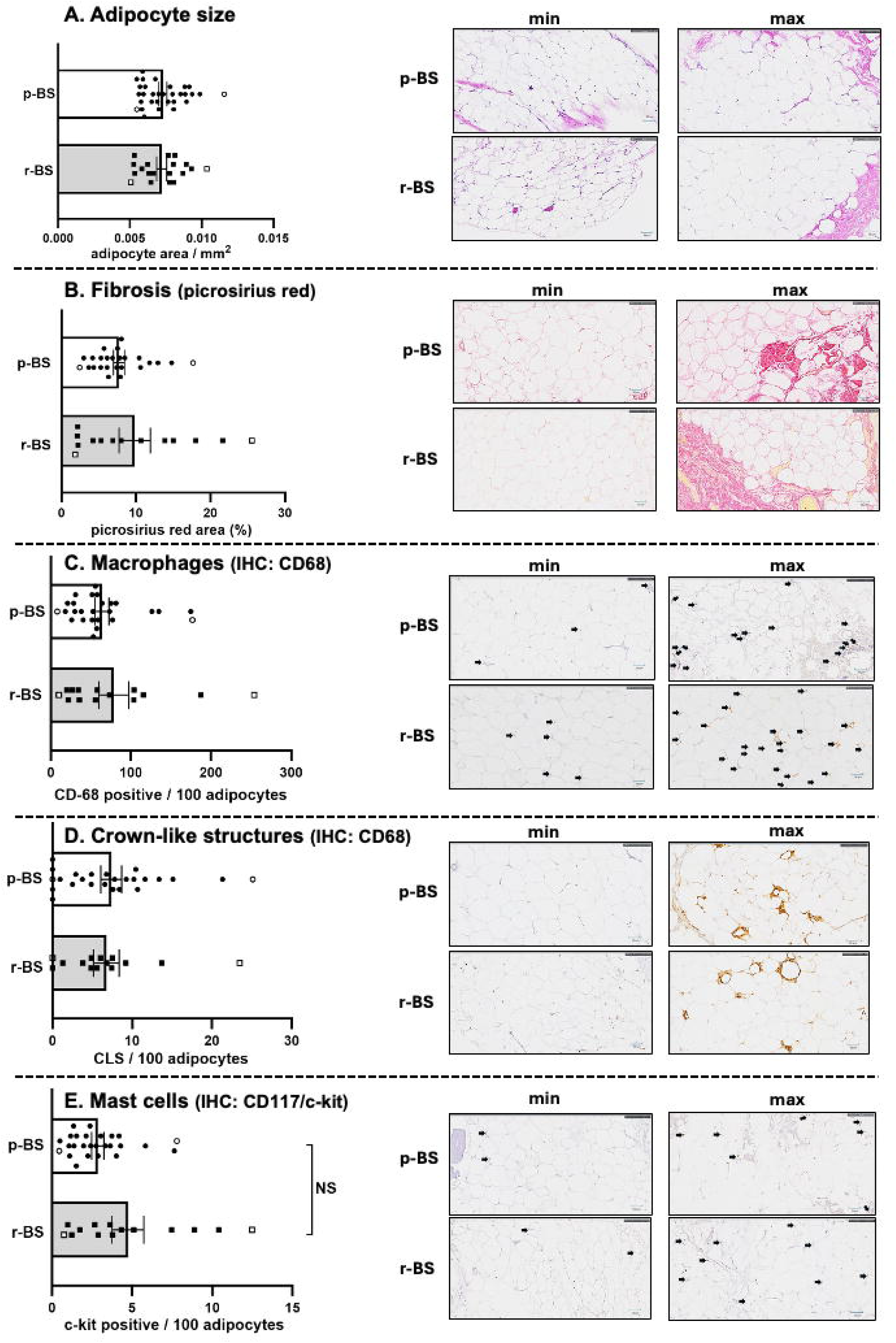
Histological analysis of abdominal subcutaneous adipose tissue samples from women with primary versus repeated bariatric surgery. Abdominal subcutaneous adipose tissues were assessed histologically, as detailed in Methods, for adipocyte size estimation (**A**.), total fibrosis (Pricosirius red staining, **B**.), macrophage infiltration (CD68 immunostaining, **C**.), crown-Like structures (CLS – i.e., CD68-positive cells surrounding the majority of an adipocyte/lipid droplet perimeter, **D**.), and infiltrating adipose tissue mast cells (CD117/c-kit immunostaining, **E**.). Each parameter is shown by the individual values obtained per group (primary bariatric surgery, p-BS, or repeated bariatric surgery, r-BS), and images illustrate the minimum and maximum values observed in each group (and depicted in the graph by an open rectangle). Comparison between the two groups was performed by Mann-Whitney test.

Next, we assessed if despite the lack of clear histological differences between the groups, adipose tissue may nevertheless present a distinct molecular fingerprint in women with a history of BS-induced significant prior weight loss and regain. For this, we performed bulk RNA-seq data of subcutaneous and visceral adipose tissue samples in a sub-cohort of n=19 of the women (10 p-BS; 9-r-BS). Like the full cohort (**Table 1**), the clinical characteristics in the sub-cohort exhibited the same group trended differences (**Table S1)**. As a first approach to assess global transcriptomic differences, we utilized principal component analysis (PCA). While analysis of visceral (omental) adipose tissue (VAT) transcriptomes showed large variability in gene expression and did not reveal any differentially expressed genes (DEGs) between the two groups (**Figure S2**), the global transcriptome of abdominal subcutaneous fat (SAT) of women in the p-BS group clearly separated from that of r-BS (**Figure 2A**). Differential expression analysis identified 90 DEGs (FDR-adjusted p-value<0.05), with 68 upregulated and 22 downregulated genes in r-BS relative to p-BS (**Figure 2B–C**, the full list of DEGs is provided in **Table S2**). These genes exhibited consistent group-specific expression patterns, as visualized in the heatmap (**Figure 2B**). Among the most dominant upregulated genes in r-BS were *JUN, JUNB*, and *FOS*, components of the AP-1 transcription factor complex ^27^, the upstream MAP-kinase-kinase-kinase (MAP3K) *TPL2* previously shown to regulate adipose tissue inflammation in obesity ^28,29^, and pro-inflammatory chemokines like *CXCL8 (IL8)*(**Figure 2C**). These all suggest activation of inflammatory and stress-response transcriptional programs in subcutaneous adipose tissue in the r-BS compared to the p-BS group.

**Figure 2.**
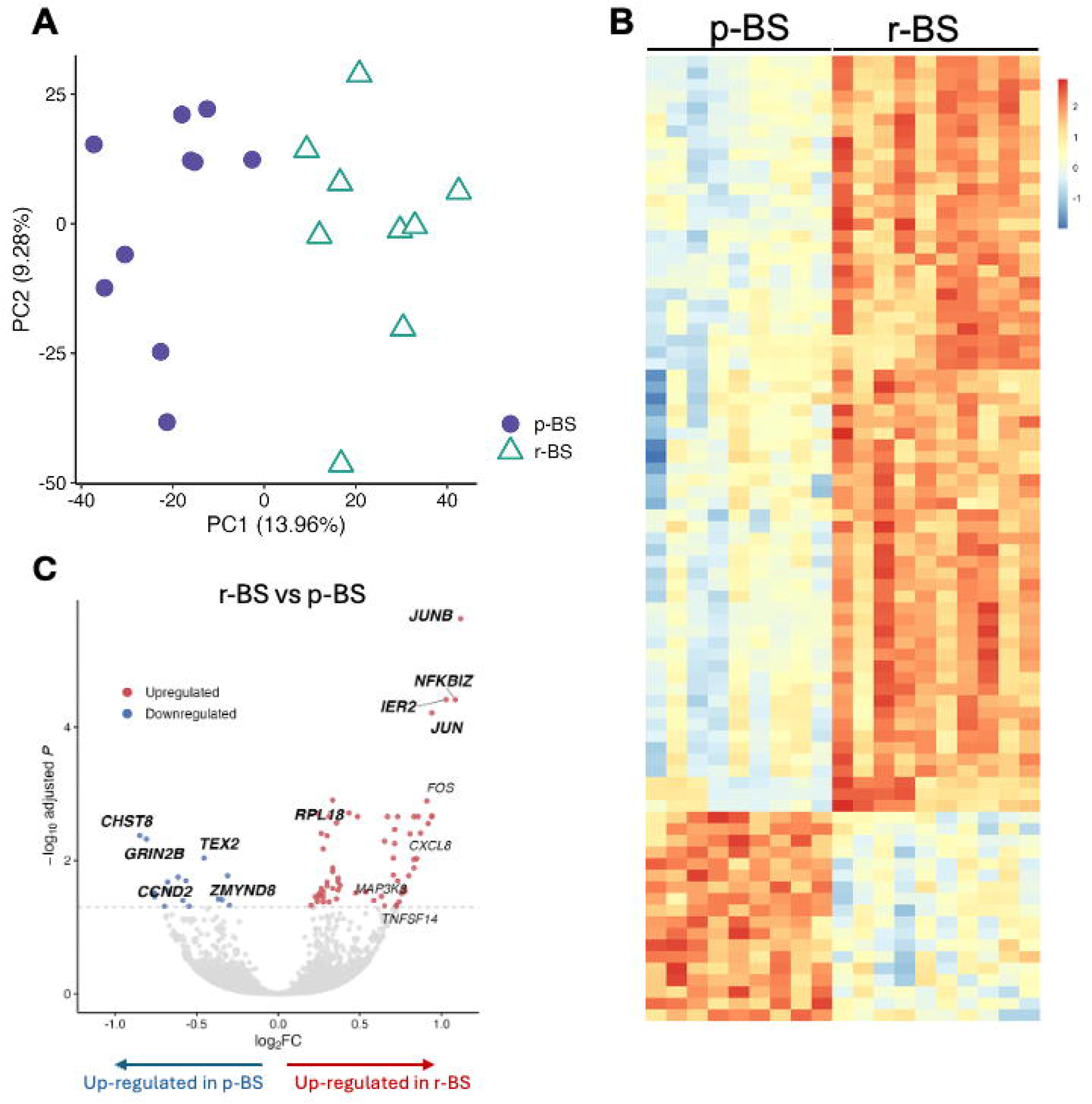
Comparison of SAT transcriptomic from women undergoing primary versus repeated bariatric surgery. **A**: Principal component analysis (PCA) of bulk RNA sequencing data from subcutaneous adipose tissue samples demonstrates separation between primary bariatric surgery (p-BS) and repeated bariatric surgery (r-BS) groups along the first principal component, indicating distinct transcriptional profiles. **B**: Heatmap of differentially expressed genes (DEGs – i.e., FDR-corrected p-value<0.05) showing relative expression (row-scaled z-scores) across samples. The full list of genes is provided in **Supplemental Table S2. C**: Volcano plot of DEGs between r-BS and p-BS. Significantly upregulated and downregulated genes (adjusted p value < 0.05) are highlighted. The top five upregulated genes in each comparative direction (i.e., p-BS vs. r-BS, and r-BS vs. p-BS), ranked by adjusted p value, are annotated (bold), along with additional selected DEGs of biological relevance. The dashed horizontal line designate the threshold FDR-adjusted p-value used to define DEGs.

To unbiasedly assess biological processes differentiating the p-BS and r-BS groups, we performed two complementary KEGG ^30^ pathway enrichment analyses: i. Over-representation analysis (ORA), which is based on genes surpassing DEG cutoff, identifying their over-representation (enrichment) in specific pathways; ii. Gene-Set Enrichment Analysis (GSEA, ^31^), which uses the entire transcriptome using ranked expression, avoiding cutoff values used to define DEGs. The genes upregulated in r-BS were enriched for ribosomal genes, and several immune-related KEGG pathways, including TNF, IL17, NF-κB, and Toll-like receptor signaling pathways (**Figure 3A**). GSEA was largely consistent with the ORA analysis. Interestingly, among the statistically significant (FDR-p<0.05) enriched pathways, those overlapping DEG-based KEGG analysis (ribosome, pro-inflammatory pathways) particularly exhibited a clear right-shift (i.e., higher expression in r-BS) of the entire set of genes forming each of the respective pathways (**Figure 3B**). Given that significant number of genes are shared by the various identified pathways, we next wished to estimate the degree of such overlap, as well as the possible transcriptional regulators of r-BS transcriptional fingerprint. Chromatin enrichment analysis (ChEA) identified multiple transcription factors that are estimated to mediate the transcriptional changes (**Table S3**), including AP-1 and NF-κB members. **Figure 3C** illustrates the joint components for the inflammatory pathways, identified by ORA, GSEA and ChEA, depicting their overlaps and unique components.

**Figure 3.**
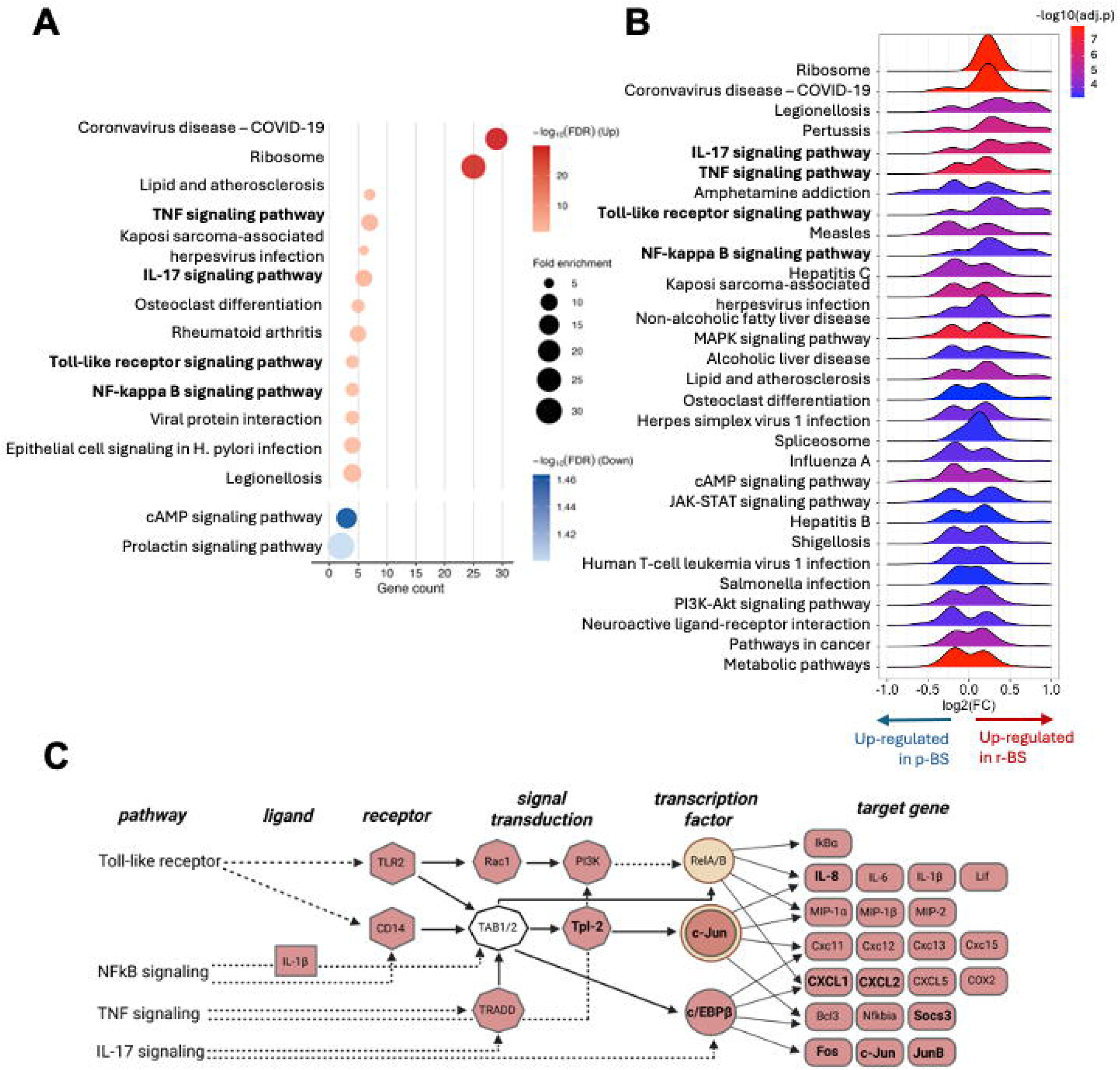
Pathway enrichment analysis in r-BS vs. p-BS comparison. **A**: DEGs were subjected to KEGG pathway over-representation enrichment analysis. The X-axis represents the number of DEGs in this pathway, dot size indicating fold enrichment, dots are colored red if up-regulated in r-BS vs. p-BS, and blue if upregulated in p-BS, and dot color intensity denotes the statistical significance of the enrichment (i.e., false discovery rate (FDR)-adjusted p). Immune and inflammatory pathways, including TNF signaling, NF-κB signaling, IL-17 signaling and Toll-like receptor signaling, are among the top enriched pathways, are shown in bold font. **B**: Gene set enrichment analysis (GSEA) ridge plot of KEGG pathways comparing r-BS to p-BS. Each ridge represents the distribution of log2 fold changes of leading-edge genes within a pathway. Ridge color corresponds to pathway enrichment significance, represented as −log10(adjusted p value). **C**: Illustration of identified joint and unique genes in the 4 inflammatory and immune-related pathways – TNF signaling, IL17 signaling, NFkB signaling and Toll-like receptor signaling. Bold – DEGs identified by KEGG (A). Non-bold are genes that belong to the leading edge in GSEA analysis (B). Red background defines up-regulated genes in r-BS compared to p-BS. Yellow designates transcription factors also identified by ChEA.

We next used a computational approach to estimate if the enrichment in inflammatory/immune pathways could reflect alterations in the cellular composition of the adipose tissue. For this we used sNucConv – a deconvolution algorithm developed based on same human adipose tissue samples sequenced both by whole-tissue (bulk) and single-nucleus RNA-sequencing to optimize the accuracy of the computational tool in estimating cellular composition from bulk RNA-seq data ^25^. Estimated cell-type proportions did not detect a robust shift in immune cell composition of SAT from p-BS versus r-BS (**Figure S3**), consistent with similar average numbers of adipose tissue macrophages and mast cells assessed histologically (**Figure 1**).

Jointly, these findings indicate that SAT obtained from women undergoing r-BS after significant weight loss and regain is characterized by a distinct transcriptional program dominated by immune and inflammatory signaling. Interestingly, this is observed without a corresponding difference in histo-pathological characteristics of the tissue, and despite the r-BS group trending to be, on one hand older, but with lower BMI and somewhat better systemic cardiometabolic risk parameters.

Since the seemingly clear pro-inflammatory transcriptional signature of SAT in r-BS corresponded with higher age, but not with a worse clinical profile, we questioned whether these changes at the mRNA levels translated functionally. To address this, we measured the secretion, at protein level, of prototypic pro-inflammatory cytokines, in conditioned media collected from SAT explants *ex-vivo* (**Figure 4A**). Adipose tissue secretion of the classical NFκB target cytokines – IL6 and IL8 (CXCL8) was indeed higher on average in conditioned media collected from r-BS, compared to p-BS, SAT samples (**Figure 4B,C**). CCL2, regulated by the up-regulated gene *NFKBIZ* (**Figure 2C**), did not exhibit a statistically significant difference (**Figure 4D**), consistent with lack of higher macrophage infiltration into the tissue (**Figure 1C**). Nevertheless, these results demonstrate that the proinflammatory transcriptional signature of r-BS SAT translates into a more proinflammatory secretory profile. To assess whether such a secretome can functionally induce inflammatory activation of other cells, we utilized human microglia HMC3 cells (**Figure 5A**). This human cell line model of “brain macrophages” responds to low concentrations of classical innate immune inflammatory activator LPS with up-regulated expression of proinflammatory genes (*IL6, TNF*) and with greater secretion of IL6 protein to the medium (**Figure 5B,C**). Given the large variation in inflammatory gene expression in the tissues, we generated an inflammatory-state rank to characterize SAT as being high versus low inflammation range (see Methods for detail). These helped to divide samples into “low-inflammatory SAT” and “high-inflammation SAT”. Conditioned media of low-inflammatory SAT did not significantly activate HMC3 cells, whether from r-BS or p-BS samples (**Figure S4A-B**). Yet, the SAT secretome of high-inflammation tissues induced a modest increase in HMC3 expression of *IL6, TNF* or *IL1B*, with somewhat higher effect in response to media from r-BS compared to p-BS (**Figure 5D**). Results were more robust when assessing protein secretion of IL6 by the human microglia cells, in which p-BS media induced 2.5-fold increase, and r-BS secretome a significantly higher, ~4.5-fold stimulation (**Figure 5E**).

**Figure 4:**
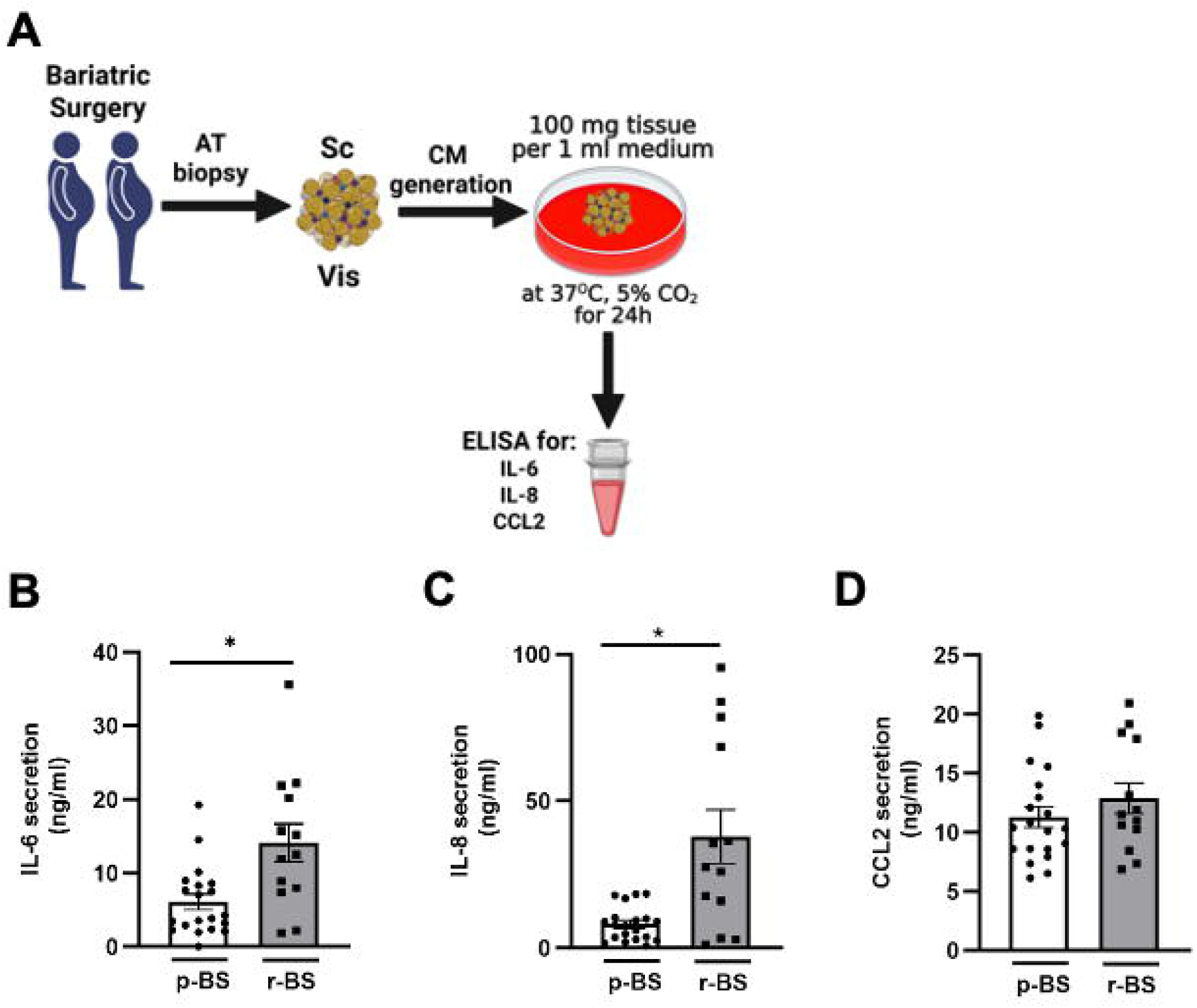
Inflammatory cytokine secretion from human subcutaneous adipose tissue explants into conditioned medium. **A**: Schematic workflow of adipose tissue conditioned medium (AT-CM) generation from biopsies of women undergoing primary versus repeated bariatric surgery (p-BS, r-BS respectively). As detailed in Methods, AT-CM was generated by incubating 100 mg tissue explants per 1 mL medium for 24 h. **B**,**C** and **D**: IL-6, IL-8 and CCL2 in AT-CM from p-BS or r-BS measured by ELISA (* p < 0.05 p-PS vs. r-BS).

**Figure 5:**
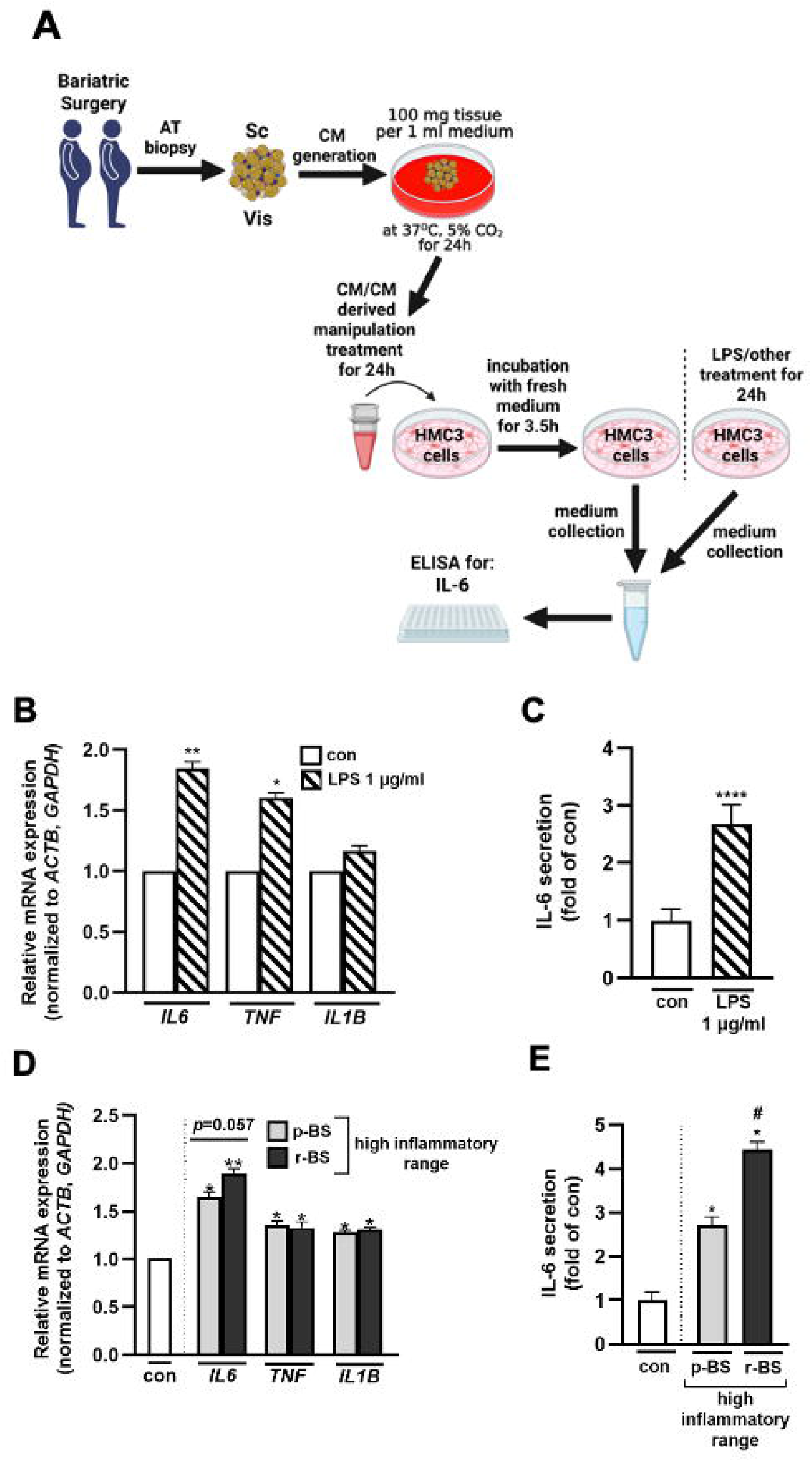
Analysis of differential activation of human microglia cells by adipose tissue conditioned media from women undergoing p-BS vs. r-BS. **A**: Schematic workflow of adipose tissue conditioned medium (AT-CM) generation from biopsies of women undergoing primary versus repeated bariatric surgery (p-BS, r-BS, respectively), and its use to treat human HMC3 microglial cells. As detailed in Methods, AT-CM was generated by incubating 100 mg tissue explants per 1 mL medium for 24 h. HMC3 cells were treated with either LPS (1 μg/ml, positive control) or AT-CM (diluted in medium 1:5) for 24 h, followed by a 3.5 h incubation in fresh medium. Cell culture supernatants were then collected for IL-6 quantification by ELISA, and cells for RNA extraction. **B**: RT-qPCR analysis of *IL6, TNF*, and *IL1B* mRNA expression (normalized to GAPDH and ACTB) in HMC3 cells stimulated with LPS (1 μg/ml) for 24 h (* p < 0.05; ** p < 0.01 relative to control). **C**: IL-6 secretion (fold-change relative to control) in HMC3 cells stimulated with LPS for 24 h, measured by ELISA. **D**: RT-qPCR analysis of *IL6, TNF*, and *IL1B* mRNA expression (normalized to GAPDH and ACTB) in HMC3 cells treated with AT-CM from p-BS or r-BS in the high-inflammatory range (* p < 0.05; ** p < 0.01 relative to control). **E**: IL-6 secretion (fold-change relative to control) in HMC3 cells treated with AT-CM from p-BS or r-BS in the high-inflammatory range, measured by ELISA (* p < 0.05 relative to control and # p < 0.05 relative to p-BS).

## Discussion

In this study we show, consistent with previous reports, that compared to p-BS candidates, r-BS women trend to clinically present as somewhat older, but with lower BMI and slightly improved cardiometabolic risk indices. This slightly distinct clinical presentation may represent an inherent (a-priori) biological difference between the two groups (that this study is blind to), or a selection bias of those undergoing r-BS. Yet, it may also reflect the beneficial effects of the significant weight loss and metabolic responses to the previous MBS persisting despite recurrent weight gain (significant weight loss after a prior MBS was an inclusion criterion for the r-BS in this study). Histological features of subcutaneous adipose tissue, including adipocyte size, fibrosis, macrophage (dispersed and in CLS) – were not different between the groups, although Mast cells trended to be more abundant in SAT of r-BS, which is consistent with their slightly better metabolic phenotype, as we previously published ^26^. Despite these, molecularly, SAT of r-BS women exhibit a clear transcriptomic fingerprint of increased inflammation. Importantly, this does not remain at the mRNA level: We demonstrate that at least some prototypic inflammatory cytokines are also secreted more from SAT of r-BS, functionally resulting in greater capacity to activate cultured human microglia cells in the subgroup of women with a high SAT inflammatory tone.

This study has several possible biases and limitations (beyond its modest cohort size), and noteworthy strengths. As mentioned above, we do not have SAT biopsies from the previous MBS of the women in the r-BS group, preventing us insight on whether the higher SAT inflammatory tone is an “inherent” a-priori biological feature of this group. This would be consistent with the proposition that higher SAT inflammatory tone contributes to the recurrent weight gain after an initial significant weight loss response to the previous MBS. Similarly, we cannot rule out clinical selection bias of r-BS candidates, or a sampling bias of SAT in patients undergoing a second laparoscopic MBS procedure. Yet, we provide an extensive histological and molecular assessment, and if the specific SAT sample was from a region affected by a tissue reaction to a prior surgical procedure, we would expect a clearer histological and/or molecular difference from p-BS, consistent with long-term response to tissue injury. Importantly, the study was restricted to female participants given limited availability of males undergoing MBS, let alone r-BS, and significant clinical and molecular differences between the sexes, consistent with known sex-specific differences in adipose tissue biology ^32^. This limits the ability to generalize our results to males. Finally, we did not identify a consistent inflammatory fingerprint in r-BS VAT due to greater variability in this tissue (from the same women) compared to SAT. Yet, our study addresses a current gap of knowledge in the understanding of differences between r-BS and p-BS at the tissue, molecular and functional levels. Importantly, we go beyond analyses of differential transcriptome: Higher expression of inflammatory pathways and mediators translate into greater secretion of inflammatory cytokines from SAT, resulting, in turn, in an inflammatory-activation of human microglia cells. While the latter setting of cultured microglia cell line exposed to conditioned medium from SAT is inherently artificial, it nevertheless demonstrates the functional potential of secreted products from high-inflammatory SAT from r-BS compared to p-BS patients.

What might be the meaning and the clinical impact of the higher SAT inflammatory state in r-BS, particularly given that their metabolic and systemic inflammatory states are not worse, and may even be somewhat better, than p-BS? Clinical studies previously reported that repeated MBS may be less effective in inducing weight loss and metabolic improvements, associated with greater risk of operative and post-operatively complications ^9,10^. Whether these can be attributed to the higher SAT inflammatory tone observed in this study is of importance, because SAT gene expression can be evaluated pre-operatively, and potentially help assess individualized MBS-related risks and outcomes. Moreover, if future studies will propose a mediatory role for SAT inflammation in poorer clinical outcomes of a repeated MBS, pre-operative intervention strategies to alleviated SAT inflammation-associated excess risk could be considered, either by immunomodulatory agents, or even PPAR-gamma and/or GLP1R agonists, which were shown to inhibit adipose tissue inflammation. Additionally, our study also suggests the potential of the secretory profile of r-BS patients to inflammatory-activate microglia – the “brain macrophages”. Obesity has been associated with neuroinflammation that could contribute to accelerated age-related cognitive decline, and possibly particularly in women – to Alzheimer’s dementia. Future studies will have to assess whether the higher inflammatory tone of SAT in r-BS candidates is connected to greater neuroinflammation and/or cognitive changes, but if so – it would again be an additional consideration for adipose inflammation targeted interventions.

In conclusion, women who undergo r-BS after an initial weight loss and recurrent weight gain present a clinically distinct subgroup of MBS candidates. Compared to those undergoing p-BS, they are likely to be older, with somewhat lower BMI, with possibly somewhat improved cardio-metabolic risk, and with a subcutaneous fat molecular and functional fingerprint consistent with higher adipose tissue inflammation.

## Data Availability

All data produced in the present study are available upon reasonable request to the authors

## Declarations

### Ethics approval and consent to participate

The study protocol was approved by the Helsinki Ethics Committee of Soroka University Medical Center (approval number: 15-0348). All participants provided written informed consent.

### Consent for publication

Not applicable.

### Availability of data and materials

The datasets generated and/or analyzed during the current study are available from the corresponding authors upon request.

### Authors’ contributions

Study conception and design – AR, YH, UY, OD; Data analysis – AS, YGN, MRL, OZ, AZ, HM; Data interpretation – AS, UY, VCC, DS, IC, RO, YH, AR; Writing the manuscript – AR, AS, YH; Critically revising the manuscript – IFL, NE, IC, RO, UY. All authors approved the final submitted version.

## Acknowledgements

The authors thank the study participants for their contribution to the Beer-Sheva adipose tissue biobank. We also thank the team of the Soroka University Medical Center Diabetes Clinic research unit, headed by Sagit Saadon and Reuma Sarousi, for their dedicated coordination and technical management of patient recruitment.

## Supplemental material

### Supplemental Figures

**Figure S1:**
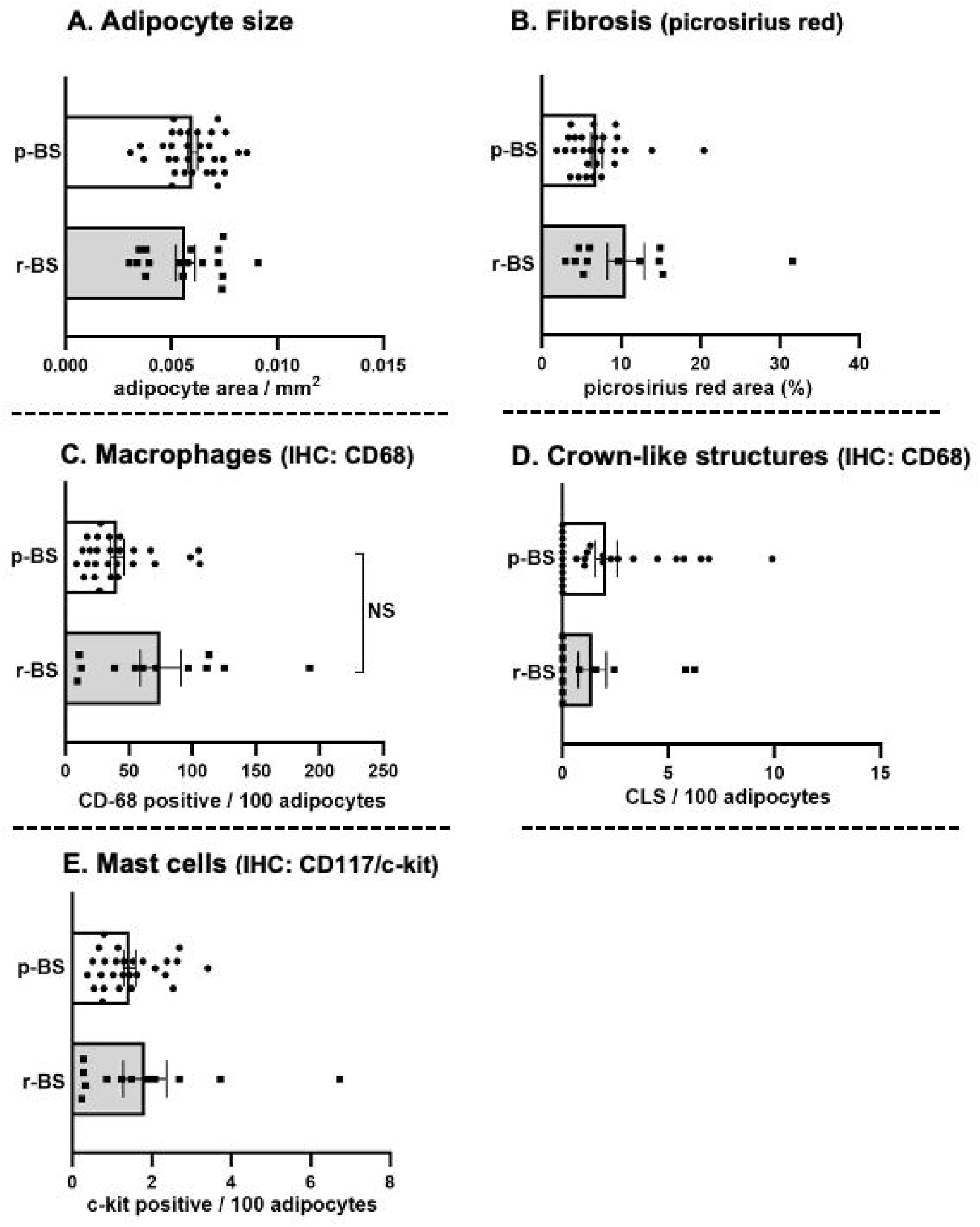
Results of histological assessment of VAT of women undergoing primary versus repeated bariatric surgery. Abdominal visceral adipose tissues were assessed histologically, as detailed in Methods, for adipocyte size estimation (**A**.), total fibrosis (Pricosirius red staining, **B**.), macrophage infiltration (CD68 immunostaining, **C**.), crown-Like structures (CLS – i.e., CD68-positive cells surrounding the majority of an adipocyte/lipid droplet perimeter, **D**.), and infiltrating adipose tissue mast cells (CD117/c-kit immunostaining, **E**.). Each parameter is shown by the individual values obtained per group (primary bariatric surgery, p-BS, or repeated bariatric surgery, r-BS). Comparison between the two groups was performed by Mann-Whitney test.

**Figure S2:**
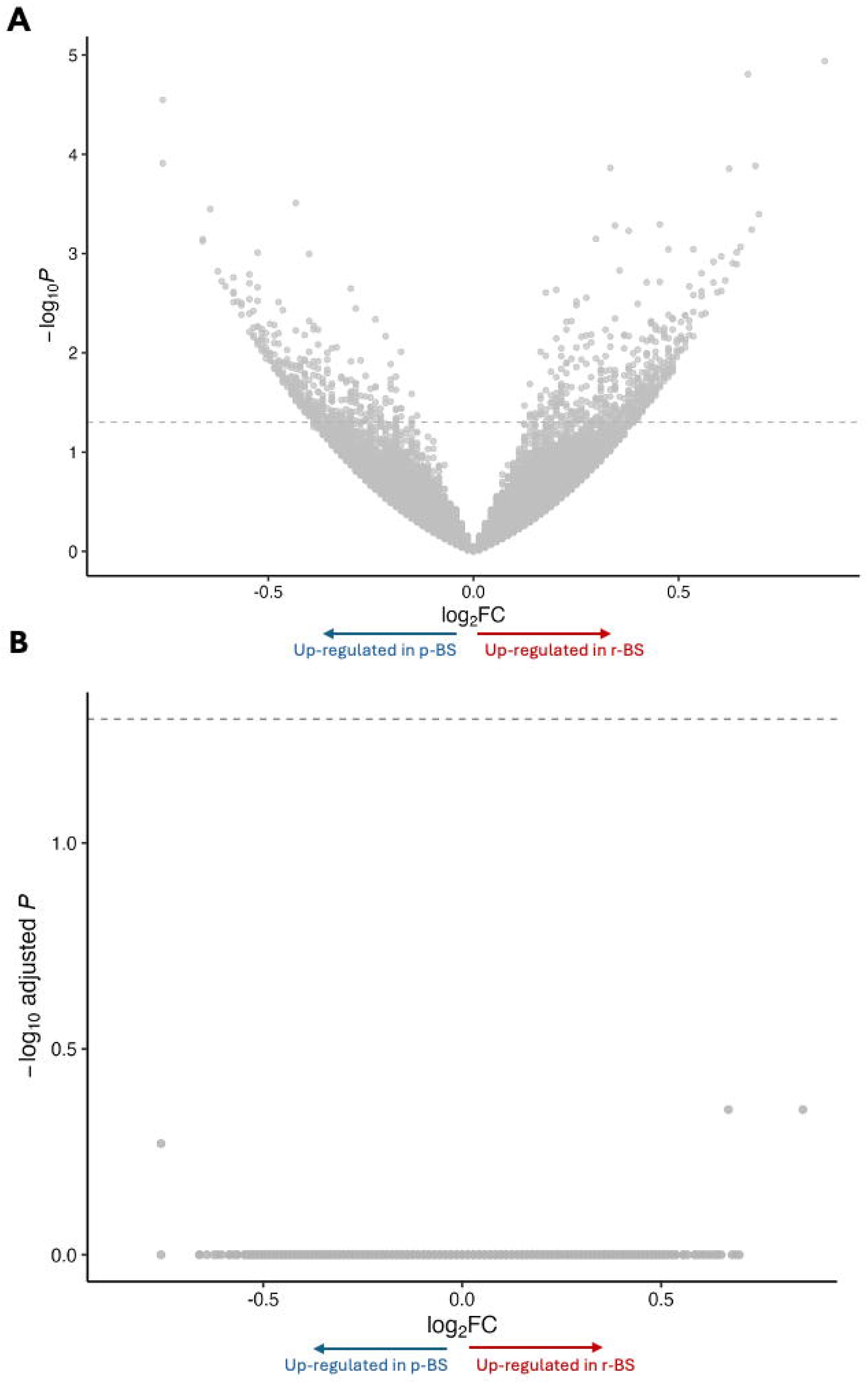
VAT RNA-seq. (A) Volcano plot of r-BS vs p-BS in VAT showing log_2_ fold change versus −log_10_(P value). Raw P values are shown for visualization. No genes were significant after multiple-testing correction. (B) Same analysis using adjusted P values (FDR), confirming no differentially expressed genes (FDR < 0.05) and lack of detectable transcriptomic differences in VAT.

**Figure S3:**
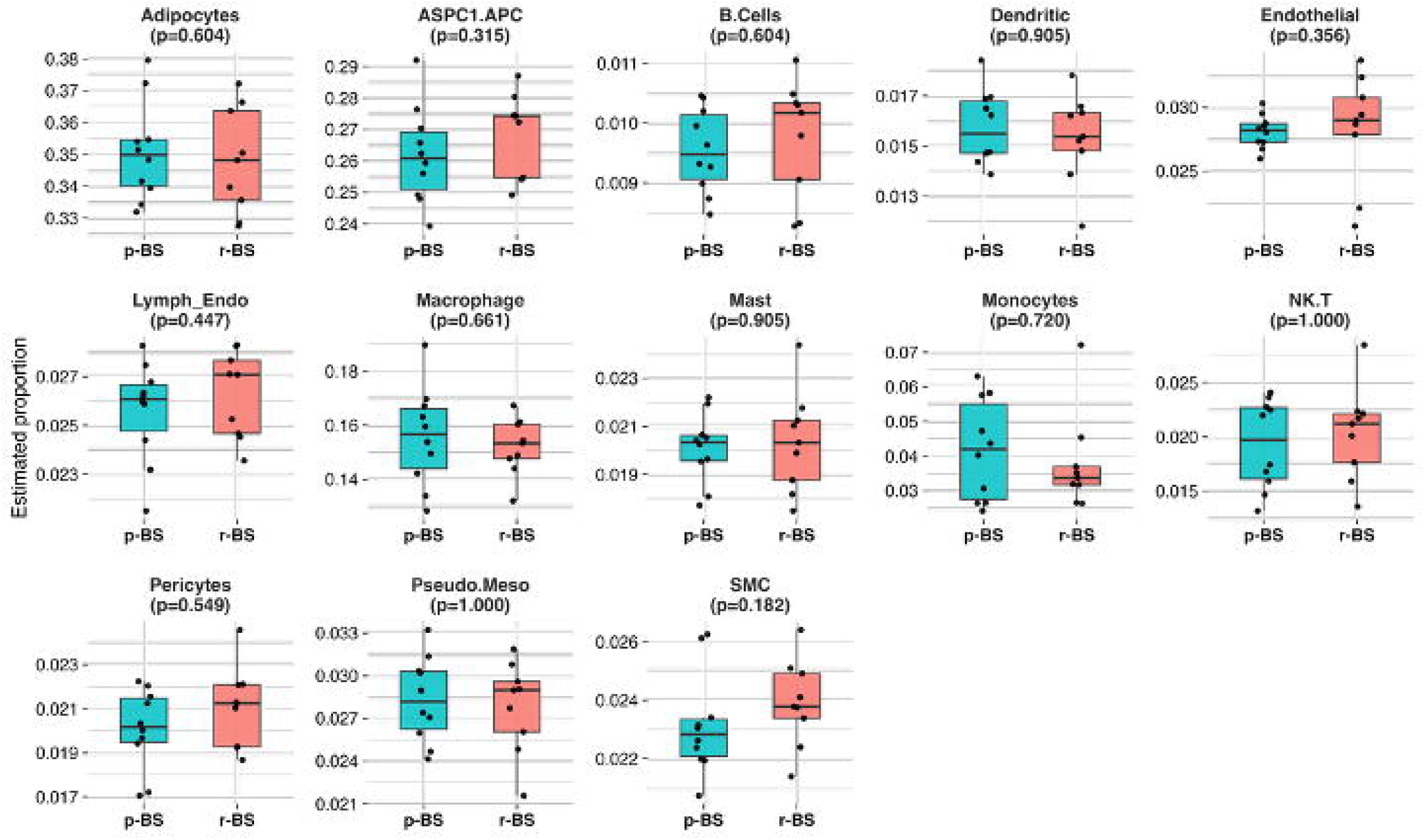
SAT cell-type composition estimated by sNucConv – a bulk-RNA-seq deconvolution algorithm. Boxplots show estimated proportions of deconvolution-derived cell types in primary bariatric surgery (p-BS) (n = 10) and repeated bariatric surgery (r-BS) (n = 9). Each panel represents a single cell type. P values (shown in panel titles) were calculated using two-tailed Mann–Whitney tests comparing p-BS and r-BS. ASPC1.APC, adipose stem and progenitor cells 1 (adipocyte progenitor cells); SMC, smooth muscle cells; NK.T, natural killer/T cells.

**Figure S4:**
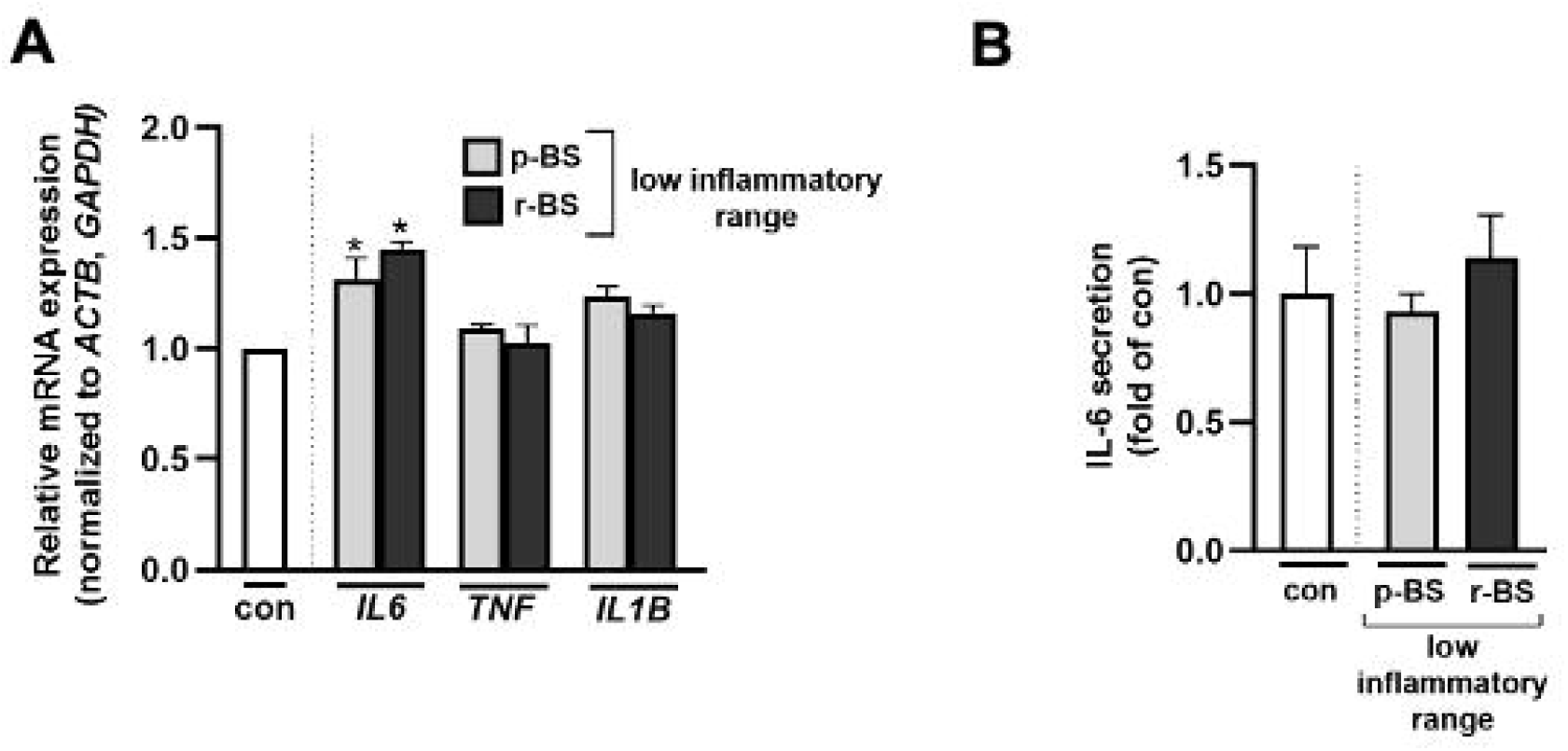
AT-CM from women undergoing p-BS vs. r-BS in the low-inflammatory range does not activate human microglia cells. **A**: RT-qPCR analysis of *IL6, TNF*, and *IL1B* mRNA expression (normalized to *GAPDH* and *ACTB*) in HMC3 cells treated with AT-CM from p-BS or r-BS in the low-inflammatory range. **B**: IL-6 secretion (fold-change relative to control) in HMC3 cells treated with AT-CM from p-BS or r-BS in the low-inflammatory range, measured by ELISA (* p < 0.05 relative to control and # p < 0.05 relative to p-BS).

### Supplemental Tables

**Table S1: Clinical characteristics of the RNA-seq sub-cohort**

**Table S2: List of differentially expressed genes in SAT**

**Table S3: List of predicted transcription factors by ChEA analysis**

## References

1. Rubino F, Nathan DM, Eckel RH, et al. Metabolic Surgery in the Treatment Algorithm for Type 2 Diabetes: A Joint Statement by International Diabetes Organizations. Diabetes Care 2016;39(6):861–77. DOI: 10.2337/dc16-0236.

2. Schauer PR, Bhatt DL, Kirwan JP, et al. Bariatric Surgery versus Intensive Medical Therapy for Diabetes - 5-Year Outcomes. N Engl J Med 2017;376(7):641–651. DOI: 10.1056/NEJMoa1600869.

3. Sjostrom L, Narbro K, Sjostrom CD, et al. Effects of bariatric surgery on mortality in Swedish obese subjects. N Engl J Med 2007;357(8):741–52. DOI: 10.1056/NEJMoa066254.

4. Angrisani L, Santonicola A, Iovino P, Formisano G, Buchwald H, Scopinaro N. Bariatric Surgery Worldwide 2013. Obes Surg 2015;25(10):1822–32. DOI: 10.1007/s11695-015-1657-z.

5. King WC, Hinerman AS, Belle SH, Wahed AS, Courcoulas AP. Comparison of the Performance of Common Measures of Weight Regain After Bariatric Surgery for Association With Clinical Outcomes. JAMA 2018;320(15):1560–1569. DOI: 10.1001/jama.2018.14433.

6. King WC, Hinerman AS, Courcoulas AP. Weight regain after bariatric surgery: a systematic literature review and comparison across studies using a large reference sample. Surg Obes Relat Dis 2020;16(8):1133–1144. DOI: 10.1016/j.soard.2020.03.034.

7. Brethauer SA, Kothari S, Sudan R, et al. Systematic review on reoperative bariatric surgery: American Society for Metabolic and Bariatric Surgery Revision Task Force. Surg Obes Relat Dis 2014;10(5):952–72. DOI: 10.1016/j.soard.2014.02.014.

8. Clapp B, Harper B, Dodoo C, et al. Trends in revisional bariatric surgery using the MBSAQIP database 2015-2017. Surg Obes Relat Dis 2020;16(7):908–915. DOI: 10.1016/j.soard.2020.03.002.

9. El Chaar M, Stoltzfus J, Melitics M, Claros L, Zeido A. 30-Day Outcomes of Revisional Bariatric Stapling Procedures: First Report Based on MBSAQIP Data Registry. Obes Surg 2018;28(8):2233–2240. DOI: 10.1007/s11695-018-3140-0.

10. Fulton C, Sheppard C, Birch D, Karmali S, de Gara C. A comparison of revisional and primary bariatric surgery. Can J Surg 2017;60(3):205–211. DOI: 10.1503/cjs.006116.

11. Giannopoulos S, Li WS, Kalantar Motamedi SM, Embry M, Stefanidis D. Outcome comparison between primary and revisional bariatric surgery: A propensity-matched analysis. Surgery 2024;175(3):592–598. DOI: 10.1016/j.surg.2023.07.027.

12. Pachter D, Klein H, Kamer O, et al. Sustained visceral fat loss is associated with attenuated brain atrophy and improved cognitive function in late midlife. Nat Commun 2026. DOI: 10.1038/s41467-026-71141-4.

13. Hinte LC, Castellano-Castillo D, Ghosh A, et al. Adipose tissue retains an epigenetic memory of obesity after weight loss. Nature 2024;636(8042):457–465. DOI: 10.1038/s41586-024-08165-7.

14. Bluher M. An overview of obesity-related complications: The epidemiological evidence linking body weight and other markers of obesity to adverse health outcomes. Diabetes Obes Metab 2025;27 Suppl 2(Suppl 2):3–19. DOI: 10.1111/dom.16263.

15. Jacks RD, Lumeng CN. Macrophage and T cell networks in adipose tissue. Nat Rev Endocrinol 2024;20(1):50–61. DOI: 10.1038/s41574-023-00908-2.

16. Sun K, Tordjman J, Clement K, Scherer PE. Fibrosis and adipose tissue dysfunction. Cell Metab 2013;18(4):470–7. DOI: 10.1016/j.cmet.2013.06.016.

17. Ouchi N, Parker JL, Lugus JJ, Walsh K. Adipokines in inflammation and metabolic disease. Nat Rev Immunol 2011;11(2):85–97. DOI: 10.1038/nri2921.

18. Liu Y, Qian SW, Tang Y, Tang QQ. The secretory function of adipose tissues in metabolic regulation. Life Metab 2024;3(2):loae003. DOI: 10.1093/lifemeta/loae003.

19. Pincu Y, Yoel U, Haim Y, et al. Assessing Obesity-Related Adipose Tissue Disease (OrAD) to Improve Precision Medicine for Patients Living With Obesity. Front Endocrinol (Lausanne) 2022;13:860799. DOI: 10.3389/fendo.2022.860799.

20. Goldstein N, Tsuneki H, Bhandarkar N, et al. Human adipose tissue is a putative direct target of daytime orexin with favorable metabolic effects: A cross-sectional study. Obesity (Silver Spring) 2021;29(11):1857–1867. DOI: 10.1002/oby.23262.

21. Maixner N, Pecht T, Haim Y, et al. A TRAIL-TL1A Paracrine Network Involving Adipocytes, Macrophages, and Lymphocytes Induces Adipose Tissue Dysfunction Downstream of E2F1 in Human Obesity. Diabetes 2020;69(11):2310–2323. DOI: 10.2337/db19-1231.

22. Pincu Y, Makarenkov N, Tsitrina AA, et al. Visceral adipocyte size links obesity with dysmetabolism more than fibrosis, and both can be estimated by circulating miRNAs. Obesity (Silver Spring) 2023;31(12):2986–2997. DOI: 10.1002/oby.23899.

23. Love MI, Huber W, Anders S. Moderated estimation of fold change and dispersion for RNA-seq data with DESeq2. Genome Biol 2014;15(12):550. DOI: 10.1186/s13059-014-0550-8.

24. Kuleshov MV, Jones MR, Rouillard AD, et al. Enrichr: a comprehensive gene set enrichment analysis web server 2016 update. Nucleic Acids Res 2016;44(W1):W90–7. DOI: 10.1093/nar/gkw377.

25. Sorek G, Haim Y, Chalifa-Caspi V, et al. sNucConv: A bulk RNA-seq deconvolution method trained on single-nucleus RNA-seq data to estimate cell-type composition of human adipose tissues. iScience 2024;27(7):110368. DOI: 10.1016/j.isci.2024.110368.

26. Goldstein N, Kezerle Y, Gepner Y, et al. Higher Mast Cell Accumulation in Human Adipose Tissues Defines Clinically Favorable Obesity Sub-Phenotypes. Cells 2020;9(6). DOI: 10.3390/cells9061508.

27. Shaulian E, Karin M. AP-1 as a regulator of cell life and death. Nat Cell Biol 2002;4(5):E131–6. DOI: 10.1038/ncb0502-e131.

28. Perfield JW, 2nd, Lee Y, Shulman GI, et al. Tumor progression locus 2 (TPL2) regulates obesity-associated inflammation and insulin resistance. Diabetes 2011;60(4):1168–76. DOI: 10.2337/db10-0715.

29. Ballak DB, van Essen P, van Diepen JA, et al. MAP3K8 (TPL2/COT) affects obesity-induced adipose tissue inflammation without systemic effects in humans and in mice. PLoS One 2014;9(2):e89615. DOI: 10.1371/journal.pone.0089615.

30. Kanehisa M, Goto S. KEGG: kyoto encyclopedia of genes and genomes. Nucleic Acids Res 2000;28(1):27–30. DOI: 10.1093/nar/28.1.27.

31. Subramanian A, Tamayo P, Mootha VK, et al. Gene set enrichment analysis: a knowledge-based approach for interpreting genome-wide expression profiles. Proc Natl Acad Sci U S A 2005;102(43):15545–50. DOI: 10.1073/pnas.0506580102.

32. Anderson WD, Soh JY, Innis SE, et al. Sex differences in human adipose tissue gene expression and genetic regulation involve adipogenesis. Genome Res 2020;30(10):1379–1392. DOI: 10.1101/gr.264614.120.

